# A Retrospective Evaluation of the Microsoft Healthcare Agent Orchestrator for Tumor Board Patient Summaries

**DOI:** 10.64898/2026.05.22.26353812

**Authors:** Joy Roy, Jack Korleski, Ryan C. Augustin, Leeor S. Yefet, Zach Jensen, Eric C. Ehman, Gelareh Zadeh, Amy Lynn Conners, Amye J. Tevaarwerk, Panagiotis Korfiatis

## Abstract

**Background:** Preparing tumor board patient summaries is time intensive. Large-language-model based systems may automate summarization but require real-world evaluation prior to clinical use. We performed an exploratory retrospective evaluation of the Microsoft Healthcare Agent Orchestrator (HAO), deployed in a Mayo Clinic–controlled staged environment, to generate tumor board–style patient summaries from retrospective Electronic Health Record (EHR) notes.

**Methods:** HAO generated summaries for breast, hepatobiliary, and neuro-oncology tumor board cases using up to the most recent 1,000 clinical notes. Clinician reviewers evaluated outputs via REDCap surveys across perceived factuality, completeness, clarity/conciseness, temporal cohesion, comparative performance, safety, and clinical utility (0–4 Likert scale). Reviewers were permitted to query the HAO chat interface to address missing details. Automated factuality was assessed using TBFact (bidirectional entailment), reporting precision and recall against available reference summaries.

**Results:** Among 57 survey responses from 5 different physicians, mean scores exceeded 2.8 across domains, with medians of 3 for most axes. In an exploratory comparison, oncology fellows required less time to review HAO-generated summaries than to manually generate patient summaries (mean difference 13.57 minutes per patient, p<0.001), although this difference may be influenced by prior familiarity with the same cases; 96% of survey responses indicated that HAO would save time. TBFact evaluations showed higher recall than precision across domains, consistent with broad capture of reference content alongside additional content that was not present in gold-standard summaries. Attribution was viewed favorably but showed issues with primary-source specificity and link reliability.

Conclusions

In a controlled Mayo environment, HAO demonstrated moderate performance and was associated with reduced review time for tumor board preparation. These findings are promising but preliminary and do not establish clinical safety, noninferiority to manual review, or readiness for routine clinical use. Limitations, including verbosity, specialty-specific content gaps, and inconsistent attribution, highlight the need for iterative refinement and further evaluation.

**Graphical Abstract:** Created using ChatGPT

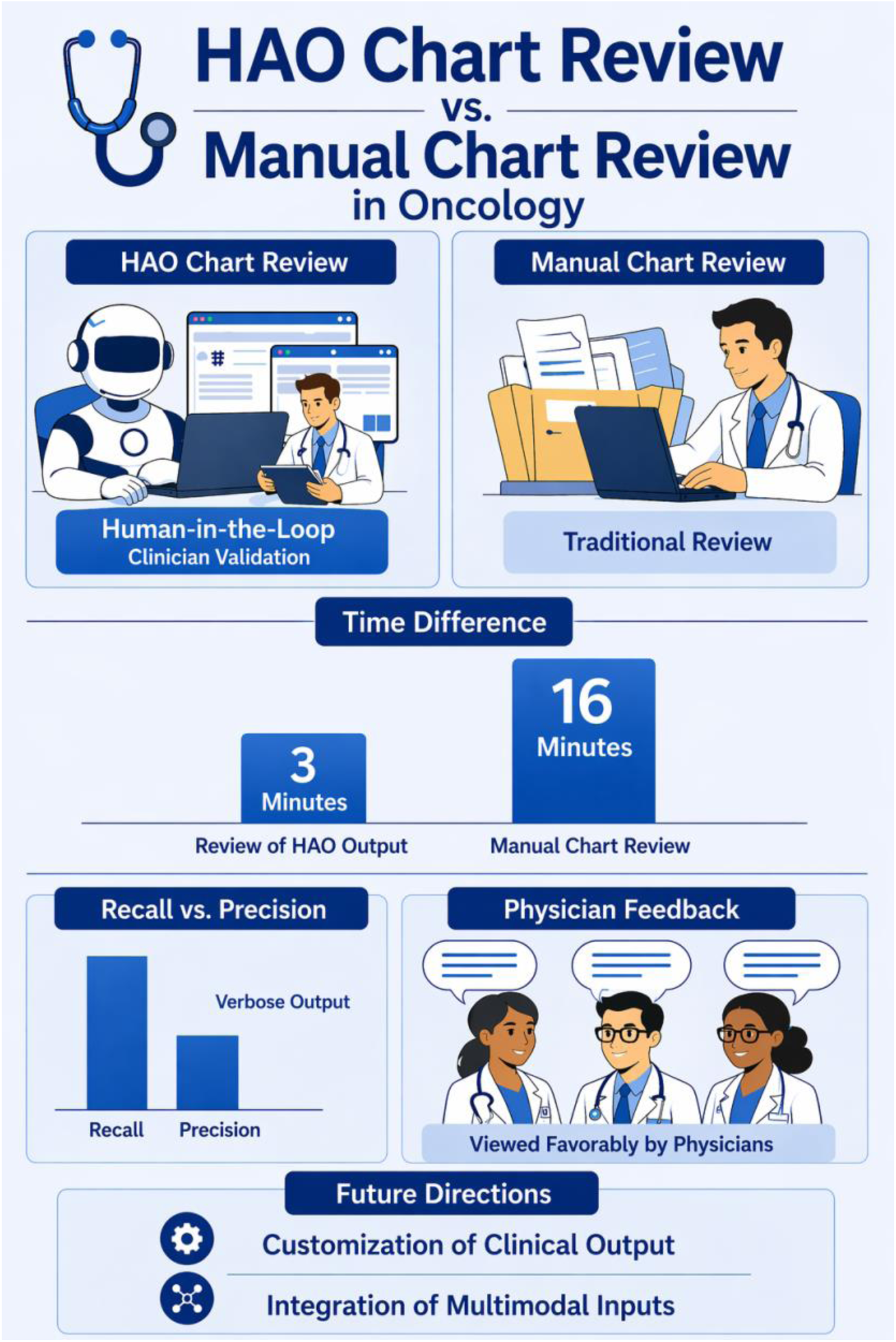

## Introduction

Multidisciplinary tumor boards (MTBs) have increasingly become the standard of care for the management of complex cancer cases ^1–6^. In these forums, specialists discuss diagnostic findings, imaging, pathology, staging, and prior treatments to collaboratively determine optimal treatment strategies^7,8^. Given high clinical workloads and the need to quickly review numerous cases, discussions are often rapid, increasing the importance of entering these meetings with well-prepared, yet concise patient histories ^9,10^. Preparing these case summaries is labor intensive and requires manual review of large volumes of electronic health record (EHR) data across fragmented systems ^11,12^. The growing number and length of clinical notes over the past decade ^13,14^ further increases the time required for comprehensive EHR review and contribute to physician burnout ^15–17^. With the rising global incidence of cancer, these challenges are likely to intensify ^18,19^.

These challenges underscore the need for innovative solutions to help reduce the time needed for EHR review while preserving clinical accuracy and safety ^20,21^. Early studies suggest that generative artificial intelligence (GenAI) assisted documentation may reduce after-hours EHR use and improve physician efficiency ^22,23^. Recently, agent-based approaches demonstrated improved performance compared with single large language models (LLM) ^24–26^.

The Microsoft Healthcare Agent Orchestrator (HAO) is a multi-agent system designed to generate structured oncology summaries by coordinating specialized agents focused on domains such as clinical history, pathology, imaging, and guideline-based reasoning ^27,28^.

Prior evaluations using curated datasets demonstrated high recall; however, as with many GenAI systems in healthcare, validation using real-world patient data and clinician assessment remains limited ^27,29–32^. Questions persist regarding factual reliability, usability, attribution quality, and clinical utility when deployed in live institutional environments with heterogeneous longitudinal documentation.

In the present study, we evaluated the HAO in a controlled Mayo Clinic environment using retrospective patient EHR data presented at MTBs across breast, hepatobiliary, and neuro-oncology specialties. Using physician evaluations collected via REDCap surveys and automated metrics, we assessed clinical utility, factual consistency, time efficiency, and limitations relevant to real-world deployment.

## Methods

### Design and Setting

We conducted a retrospective evaluation of the HAO using EHR data from patients presented at 2025 Mayo Clinic MTBs across breast, hepatobiliary, and neuro-oncology specialties. Cases were selected in collaboration with participating physicians and reflected contemporary tumor board practice. This study was approved by the Mayo Clinic Institutional Review Board (Ref #25-014848).

### Clinical data collection

Clinical notes were extracted from Mayo Clinic’s Epic EHR and converted to JSON for model ingestion. Inputs were limited to the most recent 1,000 notes to accommodate model context length constraints. Because many externally sourced documents are available only in image-based formats (e.g., scans), analyses were restricted to internal Mayo Clinic–generated clinical notes available in native text format. Data were stored in a secure Azure environment.

In addition to clinical notes, clinician-authored patient summaries were available for breast cancer and neuro-oncology. These summaries were prepared by treating clinicians for Mayo Clinic MTB presentations. Breast cancer summaries were sourced from a Mayo Clinic Health System tumor board in La Crosse, Wisconsin and are referred to as the La Crosse tumor board dataset. Neuro-oncology summaries were created using a standardized REDCap-based instrument, referred to as the Neuromets survey. Both summary types were converted to text and used as reference materials for evaluation.

### Healthcare Agent Orchestrator Implementation and Use

The HAO is a publicly available, open-source project hosted on GitHub (https://github.com/Azure-Samples/healthcare-agent-orchestrator). Microsoft representatives provided consultation and made minimal modifications to the open-source code to support deployment within the Mayo Clinic environment. Microsoft did not have access to the Mayo Clinic local codebase or any patient data throughout the study. HAO was deployed in a secure Azure environment using GPT-4.1. The HAO was not directly connected to live patient EHR records. Additional implementation details are provided in the Supplement.

### Evaluation Approach and Physician Review Workflow

To accommodate the time constraints and varying levels of engagement among participating physicians, three complementary evaluation approaches were employed. Physicians evaluated HAO-generated summaries using REDCap surveys ^33^ assessing factuality, completeness, clarity, temporal organization, safety, and clinical utility. Each domain was scored on a 5-point Likert scale ranging from 0 to 4, with 4 indicating the highest rating. Physicians also reported review time and whether HAO reduced workload. Participants were permitted to interact with the HAO to query missing information.

Physician feedback was collected using three distinct evaluation pathways:

1. **General Physician Review:** Attending-level physicians (AT, EE) reviewed the HAO-generated word document and the online HAO portal.
2. **Manual Summary Comparison by Oncology Fellows:** Two oncology fellows (RA, JK) were provided patient identifiers and manually authored their own patient summaries while timing their efforts. They subsequently reviewed the HAO-generated word document and online portal output and reported the time required to review the HAO-generated summaries.
3. **Neuromets Survey-Based Evaluation:** A study member prompted the HAO through its chat portal using structured inputs aligned with the Neuromets survey instrument to generate responses to each survey question. A neurosurgery resident (LY) then reviewed the Neuromets-style HAO output and reported feedback.

### Attribution Quality

The HAO online portal provides traceable links to source clinical notes corresponding to individual facts or summary statements. This attribution feature is available exclusively within the online chat interface and is not included in the word document. To evaluate attribution quality, a secondary REDCap survey was developed and administered to two oncology fellows (RA, JK).

### Automated Evaluation

Factual consistency was assessed using the TBFact metric, a large language model (LLM)-as-judge approach that evaluates entailment between generated summaries and reference documents ^27^. Analyses were performed across three reference sets: La Crosse breast tumor board summaries, oncology-fellow generated summaries, and Neuromets survey outputs. Results were averaged across three runs.

All primary TBFact evaluations were performed using GPT-4.1 as the LLM judge, consistent with the default HAO implementation. As an exploratory cross-model evaluation, we additionally evaluated TBFact using Gemini 2.5 Flash and Claude Haiku while preserving the same prompting framework; these results are shown in **Supplemental Figure 4**.

Cost Analysis:

Per-patient cost was estimated using Azure cost data rather than a direct per-request metric, by dividing total Azure OpenAI usage by the number of processed cases.

## Results

### HAO chart review shows moderate performance metrics

In total, we received clinical notes for 100 patients, including 50 hepatobiliary, 30 breast, and 20 neuro-oncology patients. The mean (median) number of notes per patient was 191.7 (101) with higher note volumes observed in neuro-oncology compared to hepatobiliary (p = 0.0318) **(Supplementary Figure 1)**.

We received 57 REDCap responses, including 3 from attending level physicians, 44 from oncology fellows (with self-timed review durations), 10 from a neurosurgery resident **(Table 1, Supplemental Tables 1-2)**. These responses were provided by 5 unique reviewers, including 2 attending physicians (medical oncology and radiology), 2 oncology fellows, and 1 neurosurgery resident.

**Table 1:** Tabular Summary of REDCap survey from ALL evaluators.

| **Dimension** | **N** | **Mean** | **Median** | **Std Dev** | **Min** | **Max** | **Q1** | **Q3** | **IQR** |
| --- | --- | --- | --- | --- | --- | --- | --- | --- | --- |
| Factuality | 57 | 2.88 | 3 | 0.76 | 1 | 4 | 2 | 3 | 1 |
| Completeness | 57 | 2.89 | 3 | 0.79 | 0 | 4 | 3 | 3 | 0 |
| Clarity | 57 | 2.93 | 3 | 0.59 | 2 | 4 | 3 | 3 | 0 |
| Temporal Cohesion | 57 | 3.3 | 3 | 0.76 | 1 | 4 | 3 | 4 | 1 |
| Comparative Performance | 57 | 3.07 | 3 | 0.78 | 0 | 4 | 3 | 4 | 1 |
| Safety | 57 | 3.02 | 3 | 0.79 | 1 | 4 | 3 | 4 | 1 |
| Willingness | 57 | 3.09 | 3 | 0.87 | 0 | 4 | 3 | 4 | 1 |
| Followup Questions | 45 | 3.2 | 3 | 0.94 | 0 | 4 | 3 | 4 | 1 |

Across all evaluated domains, average scores exceeded 2.8 **(Figure 2),** with a median of 3, indicating overall moderate-to-good performance metrics. When allowing the physicians to prompt the HAO further, there was not a clear improvement in completeness across all cases **(Figure 2B-C)**. In selected cases, additional prompting was associated with improved factuality **(Figure 2A)**. An example evaluation profile for a single patient (Patient 1) is shown in **Figure 2**. In this case, factuality received a score of 2, Willingness to Use a score of 3, and Completeness a score of 3; however, when reviewers were provided access to the online HAO chat portal, the Completeness score increased to 4 **(Figure 2A),** highlighting the potential of the HAO to improve with human feedback.

**Figure 1.**
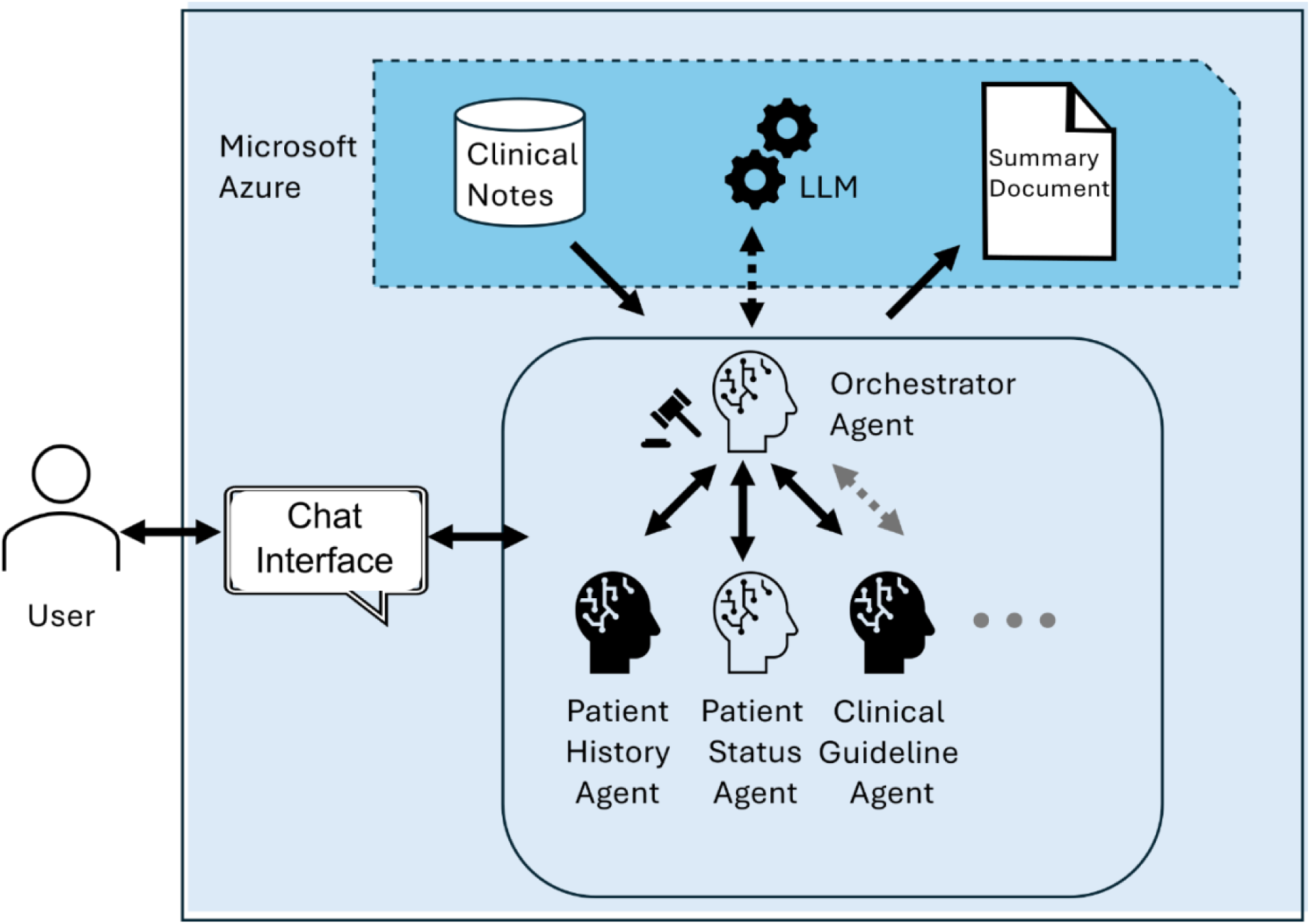
Summary schema of the HAO. The user interacts with the Microsoft HAO through a chat interface that is accessible to both the user and all individual agents. Multiple agents participate in the system and are predefined through configuration files. These agents are largely independent and extensible, allowing additional agents to be incorporated as needed. The orchestrator agent serves as the conductor of the conversation: it plans how to address the user’s request and facilitate communication among agents. Agents interact with Microsoft Azure to retrieve clinical documents and to store generated summary files. They also use Azure services, including Azure OpenAI, to invoke large language models (LLMs) for reasoning and task execution.

**Figure 2.**
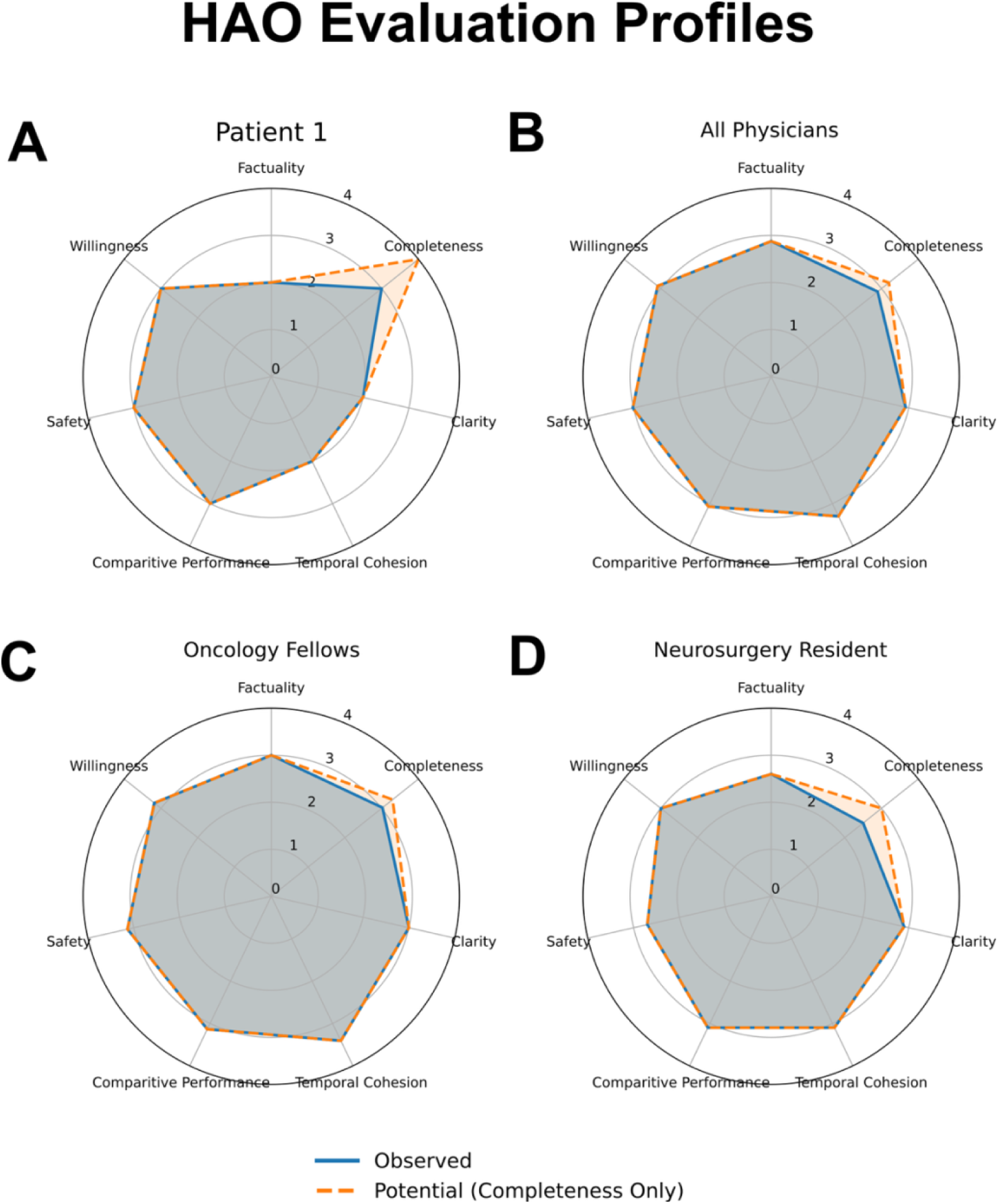
Spider plot profiles of REDCap survey results. Panel A shows responses for a single patient (“Patient 1”). Panel B displays aggregate responses from all participating physicians, Panel C displays aggregate responses from oncology fellows, and Panel D displays aggregate responses from the neurosurgery resident. All axes use a Likert scale ranging from 0 to 4, with 4 indicating the highest possible score. The shaded orange area represents scores obtained after physicians were able to query the HAO portal to address unanswered questions in the document; under this condition, only completeness was reassessed. For example, in Panel A, the default HAO summary output received a completeness score of 3, which increased to 4 (the maximum score) after physicians were able to query the HAO portal.

We next evaluated attribution quality through a separate survey due to time constraints and the large number of links provided by HAO. This survey was conducted by oncology fellows (RA, JK), who assessed attribution quality among 20 patients in the same set used for the other evaluations. On average, attribution quality received a mean score of 2.55, with a median score of 3.

### HAO Summaries demonstrated higher recall than precision

To further evaluate factuality, we conducted a TBFact analysis which reports overall precision, recall, and F1 scores, as well as category-specific scores for demographics, diagnosis, treatment, symptoms, biomarkers, and other. Because of variability in case complexity, summary length, and the number of extracted facts, we report both unweighted and weighted average precision, recall, and F1 scores. This analysis was performed across three reference sets: La Crosse breast MTB summaries **(Supplemental Table 3)**, summaries completed by oncology fellows **(Supplemental Table 4)**, and Neuromets neuro-oncology summaries **(Supplemental Table 5)**.

Across domains and scoring approaches, recall consistently exceeded precision. Compared with La Crosse MTB summaries, HAO-generated summaries achieved a mean F1 score of 0.38 (weighted 0.46). Similar performance was observed against oncology-fellow summaries (0.40 [weighted 0.49]) and neurosurgery summaries (0.35 [weighted 0.44]). Performance varied by category, with higher scores for demographics and biomarkers and lower scores for treatment.

### HAO agentic chart review was associated with reduced review time

We compared the time required to manually write patient summaries with the time required to review HAO-generated summaries for tumor board preparation. For this analysis, the oncology-fellows manually authored and timed their own patient summaries. Manual summary development required a mean of 16.52 minutes (median, 16 minutes), whereas reviewing an HAO-generated summary required a mean of 2.95 minutes (median, 3 minutes), resulting in a mean difference of 13.57 minutes per patient (p < 0.001) **(Figure 4, Supplemental Table 6)**.

Overall, in 96% of patient cases (N = 58), the respondent reported the HAO would save them time. This group included both the oncology fellows who directly timed themselves and additional physicians who responded based on clinical judgment. In contrast, our neurosurgery resident reported that HAO would not save time in 2 of the 10 cases reviewed, due to omission of key events in the clinical history.

### Latency and Cost of HAO in Tumor Board Preparation

Mean latency was approximately 3 minutes per case and appeared to increase with note length (**Supplementary Figure 2)**. 2 of 100 summaries failed due to context length limitations; both cases involved patients with particularly high note volumes indicating constraints in processing patients with extensive documentation. Per-patient cost was approximately $2–$3.

## Discussion

In this study, we evaluated both the perceived utility and current performance of the Microsoft HAO within a Mayo Clinic environment. The HAO is an open-source, multi-agent AI tool designed to ingest patient clinical notes and generate curated summaries to support MTB reviews.

### Impressions on Clinical Utility

Overall, we found that the HAO demonstrated moderate performance with some cases rated favorably compared with manual chart review. The chat-based interface enabled real-time questioning and dynamic exploration of patient history, which is not possible with a static summary document.

Additionally, the chat interface’s ability to embed links to verify source clinical notes was particularly insightful. However, limitations were identified. In some instances, HAO did not cite the original source document (e.g., pathology report) and instead referenced a secondary note. This may reflect the tool’s lack of access to original reports available only in PDF format and therefore not readily ingestible. There were also cases in which factual statements lacked appropriate citations despite warranting one, links were invalid or did not resolve correctly, and the same clinical note was repeatedly cited across multiple facts even when distinct source documents were available. Importantly, while the exclusion of PDF-only documents from the evaluation corpus may explain some failures to cite original source materials, it does not account for other issues observed, such presence of invalid source links.

### Factuality and Error Profile of HAO Summaries

Across automated evaluations, recall consistently exceeded precision, indicating that HAO captured a large proportion of clinically relevant information while also generating additional content beyond that included in reference summaries. Recall represents the proportion of reference facts captured in HAO summaries, whereas precision reflects the proportion of HAO-generated facts present in the reference summaries; higher recall therefore suggests that reference content was more frequently included in HAO outputs than the reverse. This pattern aligns with physician feedback that HAO outputs were often more comprehensive but also more verbose, consistent with prior literature that large language models often produce longer outputs in oncologic tasks ^30,34^.

Lower precision in this context does not necessarily indicate inaccuracy, but rather differences in content selection between generated and reference summaries. A key limitation of TBFact is that “not entailed” classifications may reflect contradiction or absence from the reference summary, meaning that some HAO-generated content labeled as unsupported may represent additional, rather than incorrect, information.

**Figure 3** further supports this pattern: reference content was more often directly or partially entailed by HAO outputs than vice versa. In the Reference → Model direction, “missing” indicates omissions from HAO summaries; in the Model → Reference direction, it reflects additional content not present in reference summaries. It remains unclear whether this additional content generated by HAO represents hallucinated information or factually accurate details not included in reference summaries ^35,36^. Favorable physician evaluations and the presence of source-linked citations for many generated statements provide some reassurance regarding factual grounding, although citation quality was variable. Definitive assessment of hallucination is an ongoing research topic and may require item-level verification of these additional statements against the full underlying EHR corpus rather than condensed reference summaries ^37,38^. Future work should incorporate corpus-level validation and explore retrieval-augmented generation (RAG) approaches to improve grounding and reduce unsupported content risk ^39,40^.

**Figure 3.**
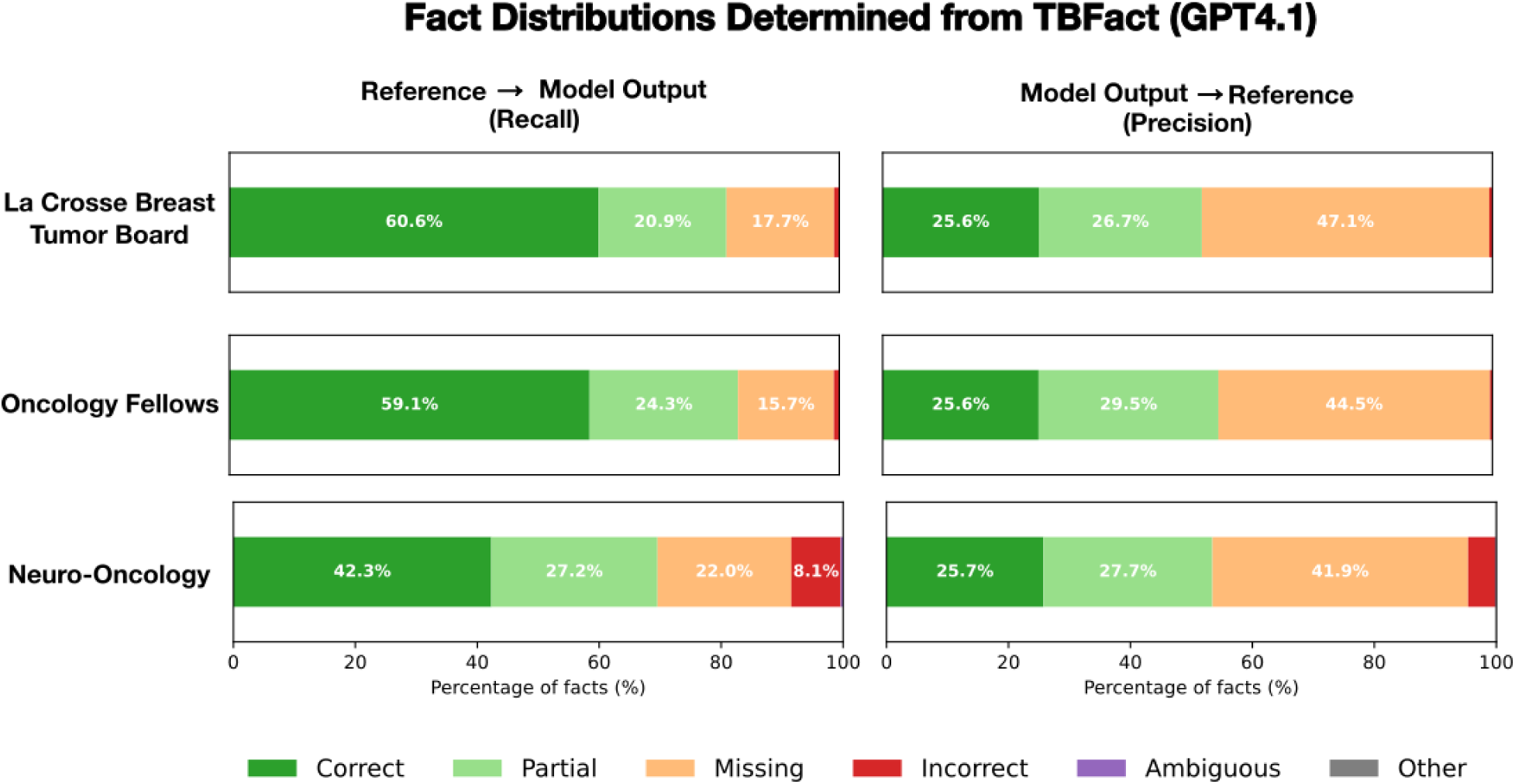
Distributions of entailment categories stratified by reference document source and direction of entailment. *Reference → Model Output* represents **recall**, assessing the proportion of facts in the reference documents that were entailed by the model output. *Model Output → Reference* represents **precision**, assessing the proportion of facts in the model output that were entailed by the reference documents. For the **La Crosse Breast dataset**, 60.6% of reference document content was fully entailed by the HAO output, 20.9% was partially entailed, 17.7% was missing, and <1% was incorrect; here, *missing* indicates content present in the reference documents but absent from the HAO output. Conversely, 25.6% of the HAO output was fully entailed by the reference documents, 26.7% was partially entailed, <1% was incorrect, and 47.1% was missing; in this direction, *missing* indicates additional content generated by HAO that was not present in the reference documents. Values shown represent the mean aggregate across three independent TBFact runs.

**Figure 4.**
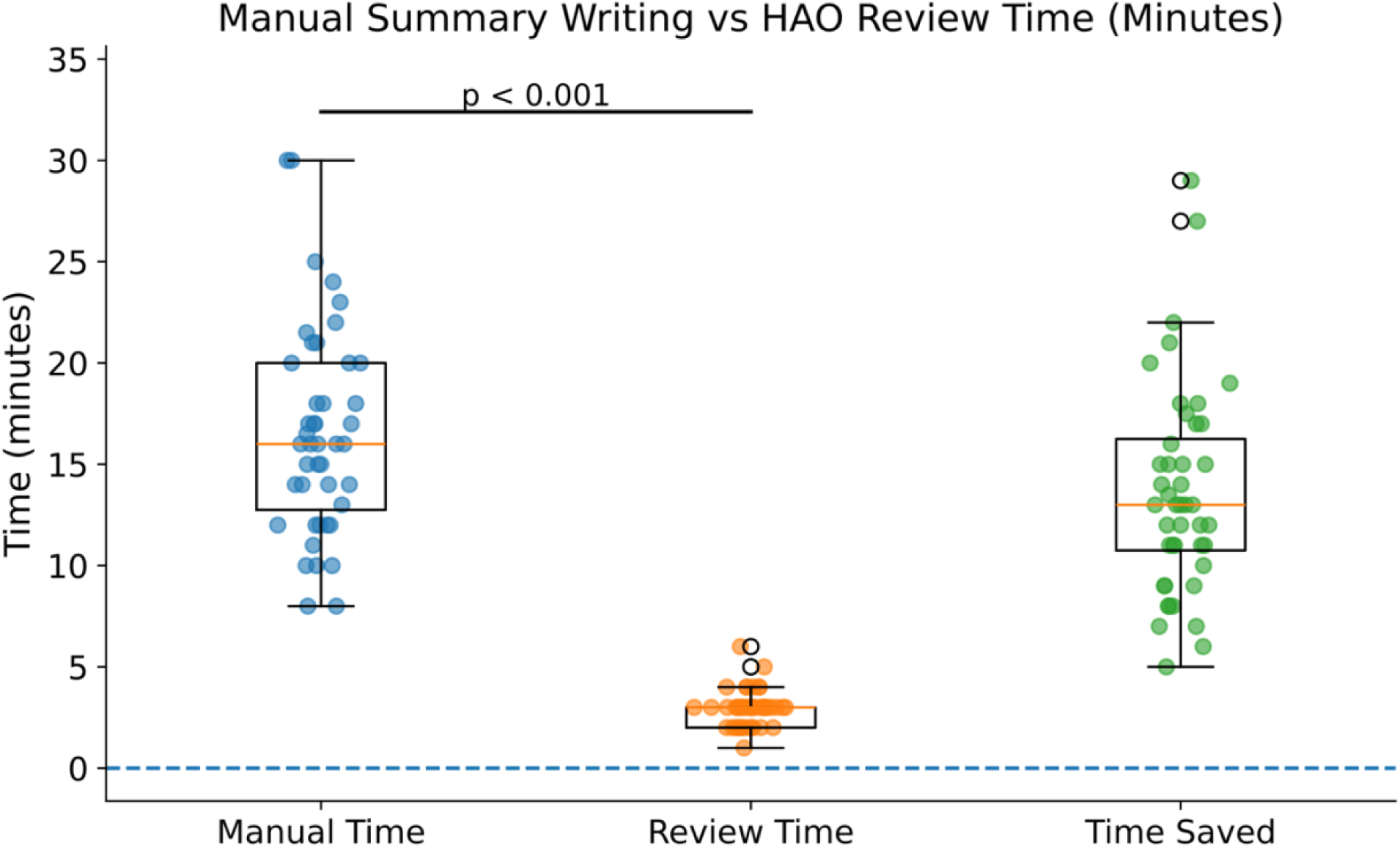
Visualization of time savings. Column 1 are the time distributions of manual preparation time needed by our oncology fellows when manually writing patient summaries. Column 2 is the time these fellows reported to review the HAO summary. Column 3 is the difference between these columns. Column 3 provides an estimate of potential time saved when only using the HAO output. The interquartile range of time savings spans from 11 to 16 min, with a median of 13 min and an average of 13.57 min.

Error rates remained low overall. For La Crosse tumor board and oncology fellow summaries, fewer than 3% of HAO-generated facts were incorrect, with ∼20% of reference content missing. In Neuro-Oncology cases, incorrect facts increased to ∼7%, with 26.5% of reference information missing and fewer directly entailed facts.

### Analysis of Review Time

The time required to review HAO-generated summaries was shorter than the time required to manually generate patient summaries, with a mean difference of 13.57 minutes per patient. However, this finding should be interpreted with caution, as the study design introduces sources of uncertainty that may influence the observed difference in either direction.

First, oncology fellows first performed manual chart review and authored patient summaries prior to reviewing the HAO-generated outputs. Prior familiarity with the patient case may therefore have reduced review times when assessing HAO summaries, potentially inflating the observed time difference. Conversely, this estimate may underestimate potential efficiency gains. The evaluation was limited to internally generated Mayo Clinic clinical notes which are generally more standardized and easier to digest than heterogeneous external records. In real-world settings, patients referred from outside institutions often require substantially more time for chart review due to variability in documentation formats and data quality^41,42^.

Taken together, these findings suggest that HAO-assisted review may reduce the time required for tumor board preparation; however, the current study design does not establish true time savings in routine clinical workflows. Future prospective evaluations, full EHR data integration will be necessary to accurately quantify the impact on clinician workload.

### Identified Limitations of the HAO

Several limitations of the HAO were identified during this evaluation. First, the system cannot currently generate dynamically formatted Word documents containing newly created content. However, HAO can be prompted to return structured outputs (e.g., JSON), which could support future dynamic document generation through downstream formatting.

Second, the default HAO output lacked certain details desired by physicians. Importantly, the specific missing details varied by clinical specialty (e.g. radiology vs medical oncology), suggesting that a single static summary format may not meet all users’ needs. Future versions of the HAO could address this limitation by incorporating specialty-specific templates to generate customized outputs.

Third, generated documents did not include links to source notes, limiting traceability and verification; adding references to static outputs would improve transparency and usability.

Fourth, HAO lacks a mechanism to filter input notes, resulting in inclusion of irrelevant data and potential context overload. Future versions should incorporate relevance-based filtering to improve efficiency and focus, as excessive or irrelevant information can impair LLM performance ^43–45^.

### Limitations of this evaluation

The HAO was used exclusively in its default configuration without prompt modification. However, this is simultaneously a strength of the study. We aimed to characterize the tool’s out-of-the-box performance, reflecting how many health care organizations would initially encounter and deploy such a system. Not all institutions will have the technical staff, resources, or governance capacity to extensively evaluate, customize, or optimize clinical AI tools prior to use ^46,47^. Key barriers to AI adoption are lack of tool maturity as well as financial concerns ^48^. Assessing default performance provides a practical baseline and informs whether further investment is warranted, while also enabling comparison with future iterations.

The clinical notes used in this evaluation were limited to internally generated Mayo Clinic documentation that were in native text format. Externally sourced records, such as scanned documents, were considered out of scope for this study and will be explored in future evaluation efforts. Notes stored in image-based formats, such as PDFs, were excluded due to reliable text extraction challenges. This constraint may also affect interpretation of both completeness and safety, as missing data types could result in omission of clinically relevant information that would be necessary for fully accurate and safe clinical decision-making. Future evaluations should consider the inclusion of optical character recognition (OCR) and related tools to enable inclusion of PDF-based documentation ^49,50^.

Safety in this study was assessed using clinician ratings rather than a direct chart-based audit; thus, findings reflect clinician-perceived safety rather than definitive clinical safety outcomes. While informative for usability and trust, these measures may not capture clinically meaningful errors or their potential impact on patient care. We did not perform systematic chart-level error review, severity grading, or dual-reviewer adjudication. Future work incorporating structured chart-based evaluation with severity classification and independent adjudication will be necessary to more rigorously assess clinical safety.

The physician survey sample was small and inter-rater reliability was not assessed. Given resource constraints, the study prioritized case diversity over agreement analysis. Future studies should include larger reviewer cohorts and formal assessment of inter-rater reliability.

## Conclusion

We present an early real-world evaluation of the Microsoft Healthcare Agent Orchestrator for generating tumor board–style summaries from EHR data. In this retrospective exploratory study, HAO generated structured summaries from diverse unstructured clinical notes with moderate overall performance and was associated with reduced review time compared with manual summary generation. These findings suggest potential utility for supporting tumor board preparation; however, they should be interpreted as preliminary. This study does not establish clinical safety, noninferiority to manual review, or readiness for routine clinical integration. Future work should prioritize improving factual accuracy, including both recall and precision, as even a small number of errors or omissions involving key clinical details could have meaningful implications for patient care and decision-making. Additional efforts should prioritize explicit signaling of missing information, improved filtering to reduce irrelevant inputs and context overload, and support for structured, template-driven outputs tailored to subspecialty needs. Subsequent evaluations should include larger sample sizes, assessment of interrater reliability, incorporation of externally sourced clinical documentation, and prospective study designs to better characterize real-world clinical impact.

## Data Availability

Data cannot be shared by the corresponding authors due to subjects’ privacy protection.

## Acknowledgements

This work is supported in part by generous funds from Dalio Philanthropies and Centurion Foundation.

## Supplement 1 – Methods

### Implementation and Reproducibility Details

The HAO is a publicly available, open-source project hosted on GitHub (https://github.com/Azure-Samples/healthcare-agent-orchestrator). The version used in this study was obtained from the repository on October 13, 2025. Microsoft representatives provided consultation and made minimal modifications to the open-source code to support deployment within the Mayo Clinic environment. Microsoft did not have access to the Mayo Clinic local codebase or any patient data at any point during the study.

HAO was deployed in a secure, Mayo Clinic–controlled Azure environment using infrastructure defined in the repository’s Bicep templates, which were used to provision the required Azure resources. The system utilized GPT-4.1 via Azure OpenAI services, selected for its larger context window after initial testing with GPT-4.o resulted in context length limitations. The HAO was not directly connected to live EHR systems. Access to the HAO chat portal was restricted to authorized users through institutional authentication and authorization controls.

#### Agent Configuration

The following agents were implemented: ‘Orchestrator’, ‘PatientHistory’, ‘PatientStatus’, ‘ClinicalGuidelines’, ‘ReportCreation’, and ‘ClinicalTrials’. However, the ‘ClinicalTrials’ agent was introduced later due to technical constraints and was not included in the evaluation. The ‘Radiology’ and ‘MedicalResearch’agents available in the repository were not used.

For TBFact evaluation, only a subset of agents (’Orchestrator’, ‘PatientHistory’, ‘PatientStatus’, and ‘ReportCreation’) was used to generate summaries, potential treatment guidelines from ‘ClinicalGuidelines’ agent and future clinical trials from ‘ClinicalTrials’agent reflect prospective recommendations that are not represented in the reference summaries.

#### User Interaction and Workflow

Users interacted with the HAO through its chat interface using standardized prompts. Specifically, users initiated summary generation with the command “@Orchestrator prepare a tumor board for patient id [id]” followed by “@Orchestrator proceed.” Upon completion, the system generated a structured Word document, which was downloaded and distributed to reviewers. Reviewers were also permitted to interact with the chat interface using additional follow-up queries.

#### Data Selection and Preprocessing

The evaluation was limited to internally generated clinical notes from the Mayo Clinic EHR. Structured data elements (e.g., laboratory values, diagnosis codes) and externally sourced or scanned documents were not included. Notes were minimally preprocessed: original text was preserved and converted into a JSON format compatible with HAO ingestion. When the number of available notes exceeded 1,000, only the most recent 1,000 notes were included to accommodate model context length limitations. This limitation only affected a single patient.

#### Case Assignment and Review Process

Patient cases were identified and then randomly assigned to reviewers by a study team member, with assignment constrained by clinical domain expertise. For example, hepatobiliary cases were reviewed by a radiologist specializing in hepatobiliary tumor boards, and neuro-oncology cases were reviewed by a neurosurgery resident. Oncology fellows reviewed cases across multiple domains, with the majority in breast oncology. Reviewers completed evaluations and REDCap surveys independently.

#### Handling of Failures

Occasional system failures occurred, typically related to session or context issues. In these cases, users were instructed to restart the session and reissue the same prompts, which generally resolved the issue. Two cases failed to complete due to context length limitations and were excluded from downstream analysis.

#### Code Modifications for Evaluation

The HAO codebase and prompting structure remained largely unchanged throughout the study. One modification was made to facilitate automated evaluation: in the default implementation, the patient timeline was embedded as an image within the Word document, which limited its usability for TBFact analysis. To address this, a study member modified the codebase to output timeline and summaries in Markdown format rather than as a Word document with embedded images. This modification was used exclusively for automated TBFact evaluation and did not affect the outputs reviewed by physicians.

### REDCaps Survey

A REDCap survey was developed to evaluate multiple dimensions of HAO performance. The following evaluation axes were assessed:

1. **Factuality**: How accurate and evidence-based is the summary?
2. **Completeness**: How fully does the summary capture the information you need?
3. **Conciseness & Clarity**: How well does the summary balance detail and readability?
4. **Temporal Cohesion**: How well are events ordered? Are they correctly labeled over time?
5. **Comparative performance**: How does the HAO summary compare to manual or existing summaries? (Skip if it doesn’t exist)
6. **Safety**: Does the summary introduce any safety-critical errors? *(Safety-critical information refers to anything that could jeopardize patient care.)*
7. **Clinical Utility**: How likely would you be to use this summary in patient care and in your workflow, either in its current form or with modest improvements?

Each domain was evaluated using a 5-point Likert scale ranging from 0 to 4, with 4 indicating the highest rating. Each Likert-scale item was followed by an open-ended question allowing respondents to explain their ratings.

Additional survey items assessed time efficiency. Physicians provided a free-text response to the question, *“How much time did you spend reviewing the patient summary?”* (excluding time spent completing the survey) and answered a binary question, *“Would this patient summary save you time?”*

Following initial testing, it became evident that the default word document output did not consistently contain all information desired by physicians. Because the evaluation focused on the overall potential of the HAO rather than its baseline static output, participants were permitted to interactively query the HAO to address unanswered questions arising from review of the document.

For the **Completeness** domain, an additional Likert-scale item was included: *“Time permitting, for missing information, does querying the HAO chatbot address the details you are missing?”*

### TBFact and Automated Evaluations

In the same GitHub repository as the HAO, Microsoft provides an automated evaluation metric called Tumor Board Fact (TBFact) ^27^. TBFact uses a large language model, in this case GPT 4.1, as its backend to assess factual consistency between texts. The metric ingests input documents, atomizes them into discrete factual statements, and evaluates entailment relationships between text bodies. It performs a bidirectional comparison between HAO-generated outputs and a gold standard reference to compute precision and recall. In addition to full entailment, TBFact also accounts for partial entailment, allowing a fact to receive partial credit when it is only partially supported by the reference text.

TBFact therefore provides a computational framework for evaluating the factuality of HAO-generated tumor board summaries. In this study, we applied TBFact using three distinct sources of gold-standard references: (1) manually written patient summaries from the Mayo Clinic La Crosse breast tumor boards; (2) manually authored summaries produced by oncology fellows during the course of this study; and (3) Neuromets surveys completed by for neuro-oncology tumor board meetings.

For evaluation purposes, we modified HAO to output summaries in Markdown format, which were then provided as input to the TBFact algorithm. We supplied only the outputs of the PatientSummary and PatientStatus agents to assess the system’s overall ability to retrieve and synthesize clinically relevant information. Given the nondeterministic nature of LLM-as-judge evaluations, all TBFact analyses were repeated three times, and aggregate results were averaged across runs. TBFact performance metrics are reported separately for each of the three reference sources.

## Supplement 2

### Summarize Physician Feedback

Several physicians provided feedback on the tool. Here, we highlight representative feedback below. Importantly, this includes both areas of strength and opportunities for improvement, as voiced by clinical users.

The Good:

- The chat-based interface is a powerful feature that allows physicians to dynamically interact with the patient record. This enables real-time questioning and exploration of patient history that is not possible with a static summary document.
- The ability to use links to verify source clinical notes was very powerful and insightful.
- The tool is most applicable and useful when evaluating patient cases that span multiple years with several clinical notes

The Bad

- Although the tool cites supporting documents for clinical facts, it does not consistently reference the original source documents (e.g., the original pathology report). Instead, it may cite downstream notes that merely restate or copy findings from the primary source.
- The default output is often too long, making it difficult to quickly extract the most relevant clinical information.
- The default output lacks sufficient radiology detail. Radiologists noted the need for structured radiology content, such as tables summarizing imaging studies, slices, and key impressions, to support efficient presentation during tumor board and multidisciplinary meetings.

The Needs (neutral and reasonably attainable)

- Clear differentiation between missing and absent data. Physicians want explicit indicators when information is unknown or not available, rather than having it silently omitted.
- Additional information including when the tumor board summary was last updated, key dates, and which physicians were responsible for specific clinical decisions or documentation.
- Customizable summaries tailored to physician specialty. Different specialties require different clinical details, and a one-size-fits-all summary does not adequately meet these varied needs.
- Ability to integrate clinical data from multiple sources and differing data types (pdf, images, unstructured text documents, etc.)

## Supplement Figures and Tables

**Supplemental Figure 1:**
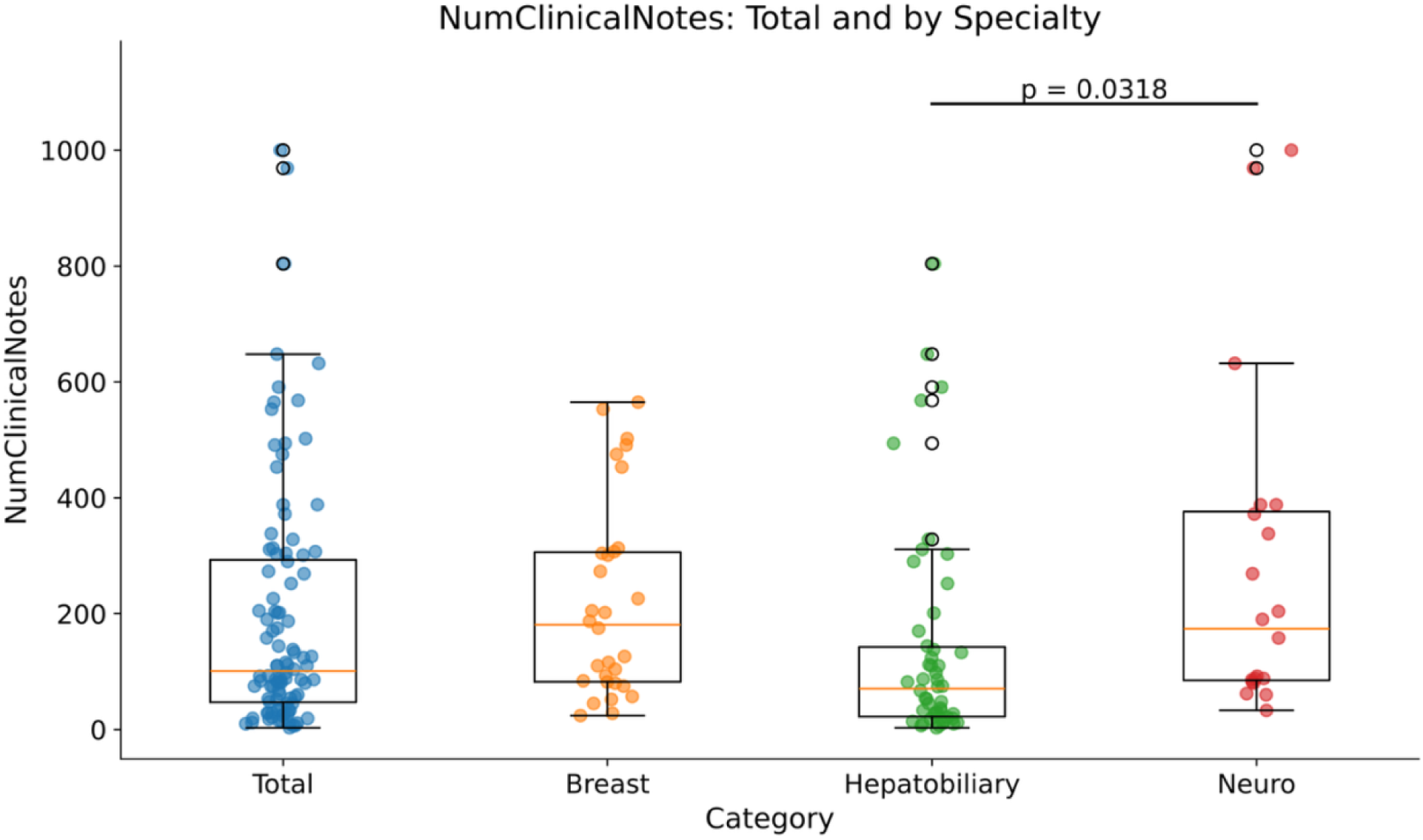
Number of clinical notes per patient by specialty. Neuro-oncology patients had significantly more clinical notes than hepatobiliary patients (p = 0.0318), with no significant difference compared with breast patients.

**Supplemental Figure 2.**
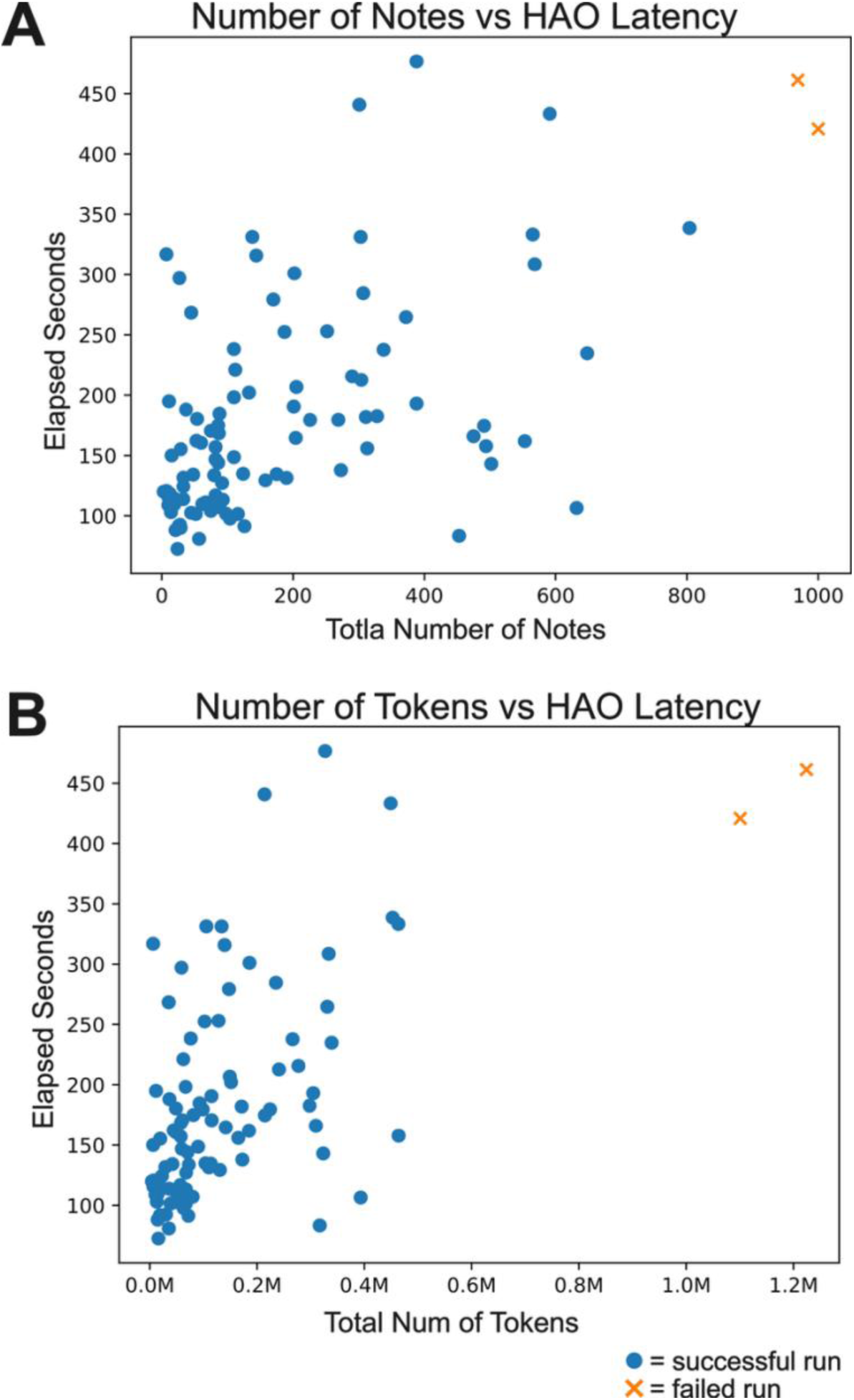
(A) Relationship between the number of clinical notes and the time required for HAO to generate a Word document. (B) The same relationship plotted against total token count. Blue circles represent successful runs that resulted in a completed Word document, whereas orange X markers indicate failed runs that did not complete due to exceeding the token limit. The average time to completion across all runs was 180 seconds.

**Supplemental Figure 3.**
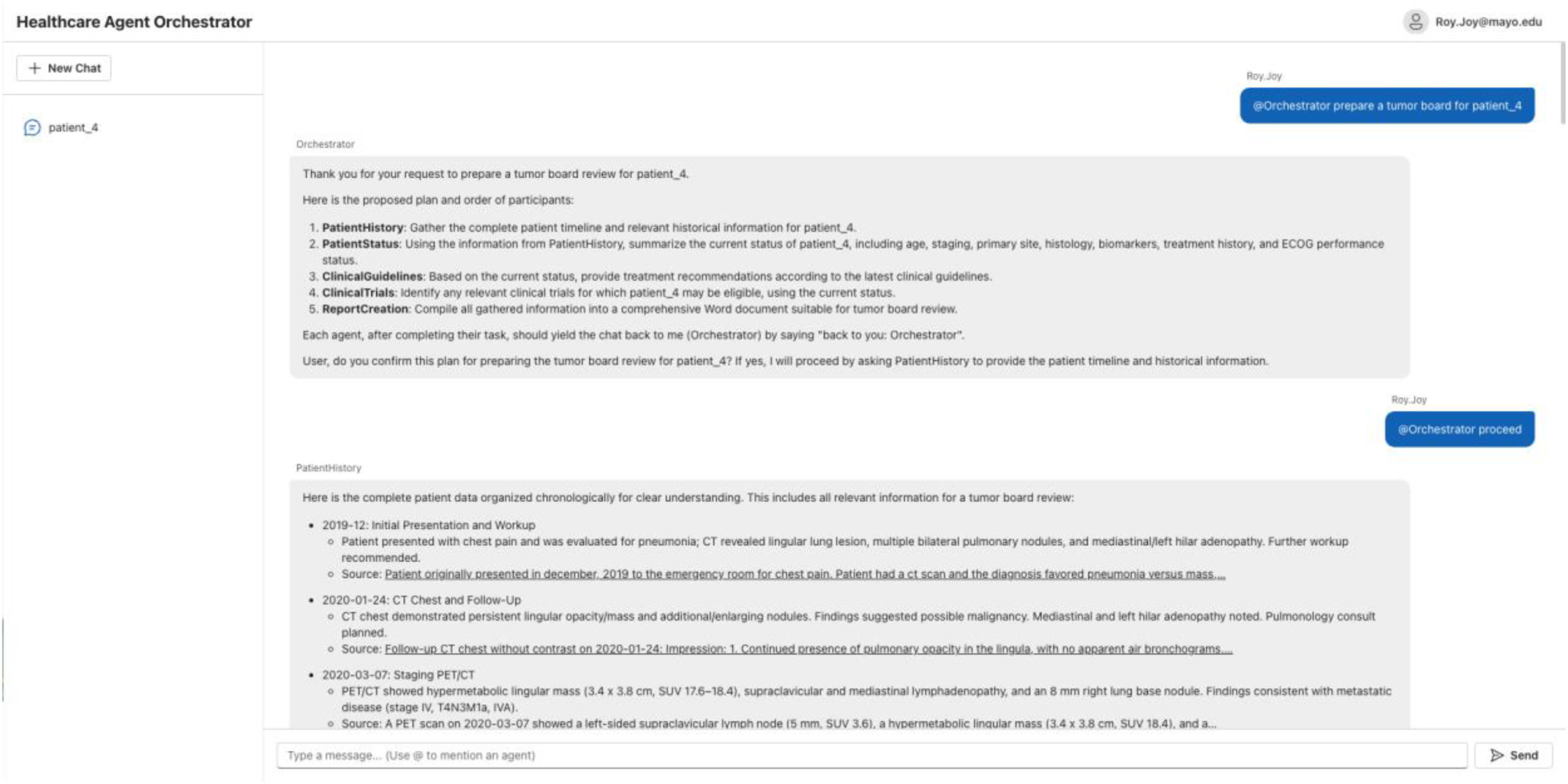
A screenshot view of the HAO Chat portal. The default view, not connected to Teams, is a chat window where you can type in your request and can even use the @ operator to query a specific agent, such as the Orchestrator agent. The following screenshot shows the interaction between user and system for a fictitious patient_4 found in the HAO Github repo.

**Supplemental Figure 4.**
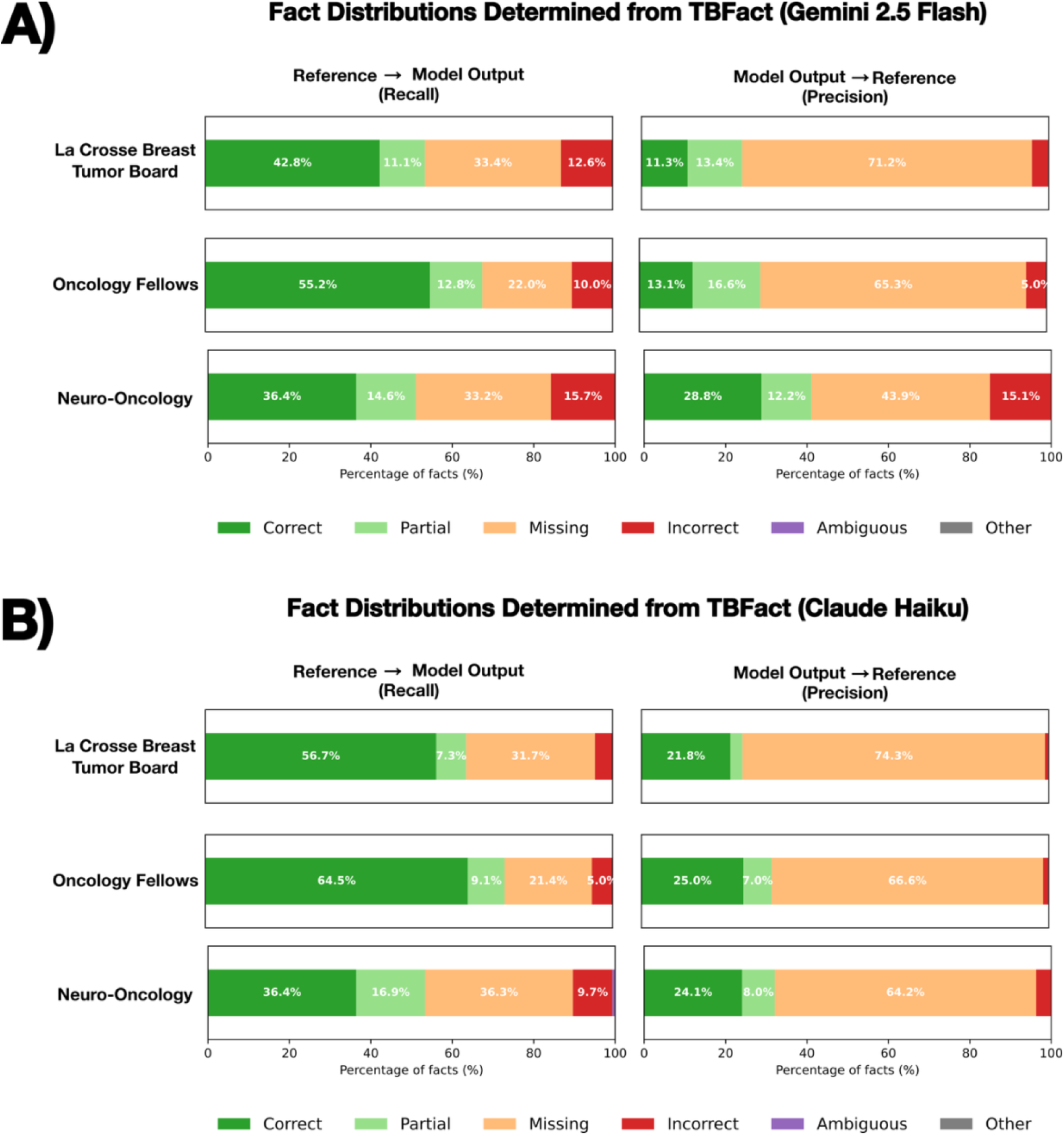
Distribution of facts identified using TBFact with (A) Gemini 2.5 Flash and (B) Claude Haiku. TBFact was included in the original HAO codebase with support for GPT models on Azure; however, we sought to investigate how alternative foundation models evaluate generated outputs. While keeping the LLM judge prompting unchanged, we modified the codebase to integrate Gemini and Claude endpoints. Compared with GPT-4.1, Gemini and Claude acted as a stricter evaluators with more identified incorrect facts and fewer partially entailed facts. Values shown represent the mean aggregate across three independent TBFact runs.

**Supplemental Table 1.**
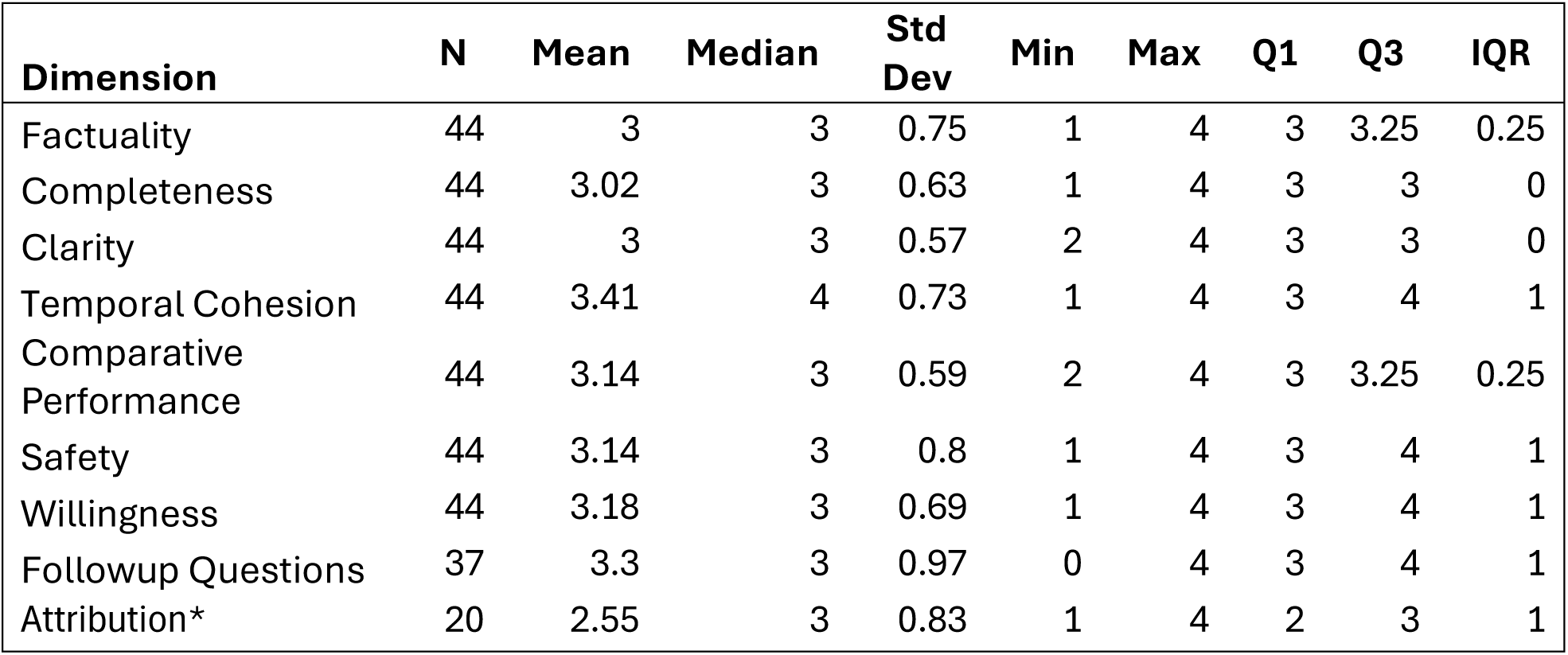
Tabulated REDCap survey results from oncology fellows (RA, JK) evaluating breast, hepatobiliary, and neuro-oncology tumor board cases alongside their corresponding HAO-generated summaries. *Attribution quality was investigated in a seperate REDCap survey only given to the oncology fellows.

| Dimension | N | Mean | Median | Std Dev | Min | Max | Q1 | Q3 | IQR |
| --- | --- | --- | --- | --- | --- | --- | --- | --- | --- |
| Factuality | 44 | 3 | 3 | 0.75 | 1 | 4 | 3 | 3.25 | 0.25 |
| Completeness | 44 | 3.02 | 3 | 0.63 | 1 | 4 | 3 | 3 | 0 |
| Clarity | 44 | 3 | 3 | 0.57 | 2 | 4 | 3 | 3 | 0 |
| Temporal Cohesion | 44 | 3.41 | 4 | 0.73 | 1 | 4 | 3 | 4 | 1 |
| Comparative Performance | 44 | 3.14 | 3 | 0.59 | 2 | 4 | 3 | 3.25 | 0.25 |
| Safety | 44 | 3.14 | 3 | 0.8 | 1 | 4 | 3 | 4 | 1 |
| Willingness | 44 | 3.18 | 3 | 0.69 | 1 | 4 | 3 | 4 | 1 |
| Followup Questions | 37 | 3.3 | 3 | 0.97 | 0 | 4 | 3 | 4 | 1 |
| Attribution* | 20 | 2.55 | 3 | 0.83 | 1 | 4 | 2 | 3 | 1 |

**Supplemental Table 2.** Tabulated REDCap survey results from neurosurgery resident evaluating neuro-oncology tumor board cases alongside their corresponding HAO-generated summaries.

| Dimension | N | Mean | Median | Std Dev | Min | Max | Q1 | Q3 | IQR |
| --- | --- | --- | --- | --- | --- | --- | --- | --- | --- |
| Factuality | 10 | 2.6 | 3 | 0.7 | 1 | 3 | 2.25 | 3 | 0.75 |
| Completeness | 10 | 2.5 | 3 | 1.27 | 0 | 4 | 2 | 3 | 1 |
| Clarity | 10 | 2.9 | 3 | 0.57 | 2 | 4 | 3 | 3 | 0 |
| Temporal Cohesion | 10 | 3.1 | 3 | 0.74 | 2 | 4 | 3 | 3.75 | 0.75 |
| Comparative Performance | 10 | 3.1 | 3.5 | 1.29 | 0 | 4 | 3 | 4 | 1 |
| Safety | 10 | 2.7 | 3 | 0.67 | 1 | 3 | 3 | 3 | 0 |
| Willingness | 10 | 3 | 3.5 | 1.41 | 0 | 4 | 3 | 4 | 1 |
| Followup Questions | 5 | 3 | 3 | 0.71 | 2 | 4 | 3 | 3 | 0 |

**Supplemental Table 3:** Using TBFact to evaluate the factuality of HAO-generated summaries against **Lacrosse Tumor Board physician–written summaries**, TBFact reported a mean F1 of 0.38 and a weighted mean F1 of 0.46. Overall, recall was generally higher than precision across domains. Values shown represent patient-level aggregates, averaged across three independent TBFact runs.

| Domain | Mean Precision | Mean Recall | Mean F1 | Weighted Mean Precision | Weighted Mean Recall | Weighted Mean F1 |
| --- | --- | --- | --- | --- | --- | --- |
| Biomarker | 0.68 | 0.72 | 0.59 | 0.69 | 0.83 | 0.75 |
| Demographics | 0.71 | 0.74 | 0.64 | 0.68 | 0.82 | 0.74 |
| Diagnosis | 0.54 | 0.67 | 0.54 | 0.55 | 0.73 | 0.63 |
| Other | 0.15 | 0.43 | 0.14 | 0.18 | 0.63 | 0.27 |
| Symptom | 0.39 | 0.64 | 0.43 | 0.39 | 0.71 | 0.5 |
| Treatment | 0.16 | 0.23 | 0.08 | 0.16 | 0.62 | 0.24 |
| Overall | 0.4 | 0.54 | 0.38 | 0.38 | 0.6 | 0.46 |

**Supplemental Table 4:** Using TBFact to evaluate the factuality of HAO-generated summaries against **Oncology fellow–written summaries**, TBFact reported a mean F1 of 0.4 with a weighted mean F1 of 0.49. Overall, recall was generally higher than precision across domains. Values shown represent patient-level aggregates, averaged across three independent TBFact runs.

| Domain | Mean Precision | Mean Recall | Mean F1 | Weighted Mean Precision | Weighted Mean Recall | Weighted Mean F1 |
| --- | --- | --- | --- | --- | --- | --- |
| Biomarker | 0.6 | 0.67 | 0.53 | 0.58 | 0.75 | 0.65 |
| Demographics | 0.69 | 0.73 | 0.64 | 0.63 | 0.82 | 0.71 |
| Diagnosis | 0.55 | 0.68 | 0.55 | 0.56 | 0.74 | 0.64 |
| Other | 0.17 | 0.41 | 0.13 | 0.18 | 0.6 | 0.28 |
| Symptom | 0.4 | 0.64 | 0.45 | 0.4 | 0.71 | 0.52 |
| Treatment | 0.21 | 0.3 | 0.15 | 0.22 | 0.66 | 0.33 |
| Overall | 0.39 | 0.58 | 0.4 | 0.39 | 0.67 | 0.49 |

**Supplemental Table 5:**
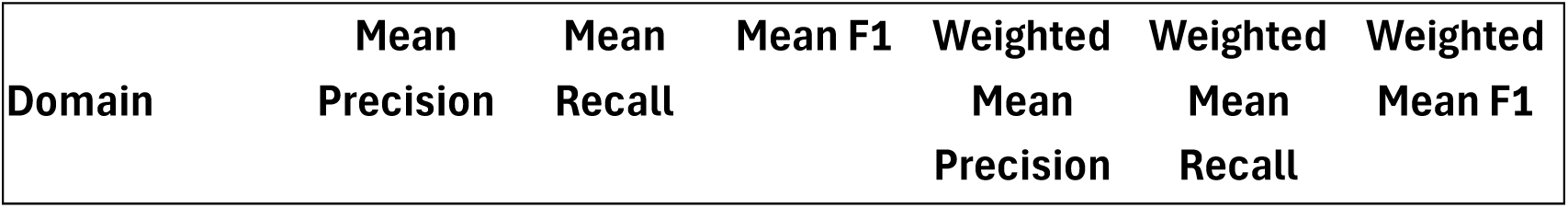
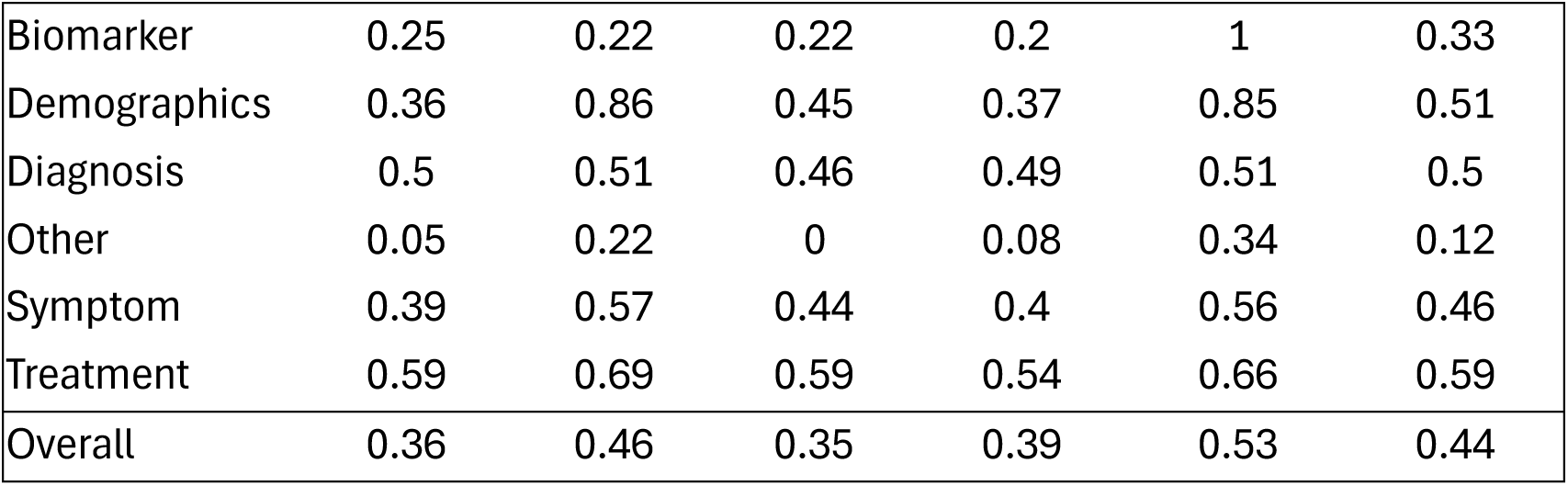
Using TBFact to evaluate the factuality of HAO-generated summaries against **neurosugery–written summaries**, TBFact reported a mean F1 of 0.35 with a weighted mean F1 of 0.44. Overall, recall was generally higher than precision across domains. Values shown represent patient-level aggregates, averaged across three independent TBFact runs.

| Domain | Mean Precision | Mean Recall | Mean F1 | Weighted Mean Precision | Weighted Mean Recall | Weighted Mean F1 |
| --- | --- | --- | --- | --- | --- | --- |
| Biomarker | 0.25 | 0.22 | 0.22 | 0.2 | 1 | 0.33 |
| Demographics | 0.36 | 0.86 | 0.45 | 0.37 | 0.85 | 0.51 |
| Diagnosis | 0.5 | 0.51 | 0.46 | 0.49 | 0.51 | 0.5 |
| Other | 0.05 | 0.22 | 0 | 0.08 | 0.34 | 0.12 |
| Symptom | 0.39 | 0.57 | 0.44 | 0.4 | 0.56 | 0.46 |
| Treatment | 0.59 | 0.69 | 0.59 | 0.54 | 0.66 | 0.59 |
| Overall | 0.36 | 0.46 | 0.35 | 0.39 | 0.53 | 0.44 |

**Supplemental Table 6.** Time Savings. Mean difference.

| **Metric** | **N** | **Mean**<br>**(min)** | **Median**<br>**(min)** | **Q1 (min)** | **Q3**<br>**(min)** | **Min**<br>**(min)** | **Max**<br>**(min)** |
| --- | --- | --- | --- | --- | --- | --- | --- |
| Manual Time | 44 | 16.52 | 16 | 12.75 | 20 | 8 | 30 |
| Review Time | 44 | 2.95 | 3 | 2 | 3 | 1 | 6 |
| Difference* | 44 | 13.57 | 13 | 10.75 | 16.25 | 5 | 29 |

\*p < 0.001

## Bibliography

1. Lutz S, D’Angelo A, Hammerl S, et al. Unveiling the Digital Evolution of Molecular Tumor Boards. Target Oncol. Jan 2025;20(1):27–43. doi:10.1007/s11523-024-01109-1

2. Specchia ML, Frisicale EM, Carini E, et al. The impact of tumor board on cancer care: evidence from an umbrella review. BMC Health Serv Res. Jan 31 2020;20(1):73. doi:10.1186/s12913-020-4930-3

3. Thenappan A, Halaweish I, Mody RJ, et al. Review at a multidisciplinary tumor board impacts critical management decisions of pediatric patients with cancer. Pediatr Blood Cancer. Feb 2017;64(2):254–258. doi:10.1002/pbc.26201

4. Di Pilla A, Cozzolino MR, Mannocci A, et al. The Impact of Tumor Boards on Breast Cancer Care: Evidence from a Systematic Literature Review and Meta-Analysis. Int J Environ Res Public Health. Nov 14 2022;19(22)doi:10.3390/ijerph192214990

5. Liu JC, Kaplon A, Blackman E, Miyamoto C, Savior D, Ragin C. The impact of the multidisciplinary tumor board on head and neck cancer outcomes. Laryngoscope. Apr 2020;130(4):946–950. doi:10.1002/lary.28066

6. Naessens C, Laloze J, Leobon S, et al. Physician compliance with multidisciplinary tumor board recommendations for managing gynecological cancers. Future Oncol. Apr 2023;19(13):897–908. doi:10.2217/fon-2022-1183

7. Winters DA, Soukup T, Sevdalis N, Green JSA, Lamb BW. The cancer multidisciplinary team meeting: in need of change? History, challenges and future perspectives. BJU Int. Sep 2021;128(3):271–279. doi:10.1111/bju.15495

8. Shah BA, Qureshi MM, Jalisi S, et al. Analysis of decision making at a multidisciplinary head and neck tumor board incorporating evidence-based National Cancer Comprehensive Network (NCCN) guidelines. Pract Radiat Oncol. Jul–Aug 2016;6(4):248–254. doi:10.1016/j.prro.2015.11.006

9. Mnajjed L, Krempl G. Increased Efficiency of Tumor Board With Introduction of the Consensus Slate. Ear Nose Throat J. Jun 25 2025:1455613251351773. doi:10.1177/01455613251351773

10. Mullan BJ, Brown JS, Lowe D, Rogers SN, Shaw RJ. Analysis of time taken to discuss new patients with head and neck cancer in multidisciplinary team meetings. Br J Oral Maxillofac Surg. Feb 2014;52(2):128–33. doi:10.1016/j.bjoms.2013.10.001

11. Hammer RD, Fowler D, Sheets LR, Siadimas A, Guo C, Prime MS. Digital Tumor Board Solutions Have Significant Impact on Case Preparation. JCO Clin Cancer Inform. Aug 2020;4:757–768. doi:10.1200/CCI.20.00029

12. Krupinski EA, Comas M, Gallego LG, Group G. A New Software Platform to Improve Multidisciplinary Tumor Board Workflows and User Satisfaction: A Pilot Study. J Pathol Inform. 2018;9:26. doi:10.4103/jpi.jpi_16_18

13. Rule A, Bedrick S, Chiang MF, Hribar MR. Length and Redundancy of Outpatient Progress Notes Across a Decade at an Academic Medical Center. JAMA Netw Open. Jul 1 2021;4(7):e2115334. doi:10.1001/jamanetworkopen.2021.15334

14. Goldstein IH, Hwang T, Gowrisankaran S, Bales R, Chiang MF, Hribar MR. Changes in Electronic Health Record Use Time and Documentation over the Course of a Decade. Ophthalmology. Jun 2019;126(6):783–791. doi:10.1016/j.ophtha.2019.01.011

15. Eschenroeder HC, Manzione LC, Adler-Milstein J, et al. Associations of physician burnout with organizational electronic health record support and after-hours charting. J Am Med Inform Assoc. Apr 23 2021;28(5):960–966. doi:10.1093/jamia/ocab053

16. Kruse CS, Mileski M, Dray G, Johnson Z, Shaw C, Shirodkar H. Physician Burnout and the Electronic Health Record Leading Up to and During the First Year of COVID-19: Systematic Review. J Med Internet Res. Mar 31 2022;24(3):e36200. doi:10.2196/36200

17. Muhiyaddin R, Elfadl A, Mohamed E, et al. Electronic Health Records and Physician Burnout: A Scoping Review. Stud Health Technol Inform. Jan 14 2022;289:481–484. doi:10.3233/SHTI210962

18. Sung H, Ferlay J, Siegel RL, et al. Global Cancer Statistics 2020: GLOBOCAN Estimates of Incidence and Mortality Worldwide for 36 Cancers in 185 Countries. CA Cancer J Clin. May 2021;71(3):209–249. doi:10.3322/caac.21660

19. Bray F, Laversanne M, Sung H, et al. Global cancer statistics 2022: GLOBOCAN estimates of incidence and mortality worldwide for 36 cancers in 185 countries. CA Cancer J Clin. May–Jun 2024;74(3):229–263. doi:10.3322/caac.21834

20. Arafat W, Fu P, Jr., Wagner AJ, et al. Clinician Perspectives Regarding the Impact of Information Technology on Multidisciplinary Tumor Boards: A National Comprehensive Cancer Network Survey. JCO Clin Cancer Inform. Sep 2023;7:e2300056. doi:10.1200/CCI.23.00056

21. Chang LC, Kuo HC, Wang HM, et al. The Use of an Integrated Digital Tool to Improve the Efficiency of Multidisciplinary Tumor Boards-A Prospective Trial in Taiwan. Cancers (Basel). Jan 28 2025;17(3)doi:10.3390/cancers17030444

22. Liu TL, Hetherington TC, Stephens C, et al. AI-Powered Clinical Documentation and Clinicians’ Electronic Health Record Experience: A Nonrandomized Clinical Trial. JAMA Netw Open. Sep 3 2024;7(9):e2432460. doi:10.1001/jamanetworkopen.2024.32460

23. Pearlman K, Wan W, Shah S, Laiteerapong N. Use of an AI Scribe and Electronic Health Record Efficiency. JAMA Netw Open. Oct 1 2025;8(10):e2537000. doi:10.1001/jamanetworkopen.2025.37000

24. Ferber D, El Nahhas OSM, Wolflein G, et al. Development and validation of an autonomous artificial intelligence agent for clinical decision-making in oncology. Nat Cancer. Aug 2025;6(8):1337–1349. doi:10.1038/s43018-025-00991-6

25. Lee Y, Wang X, Yang CC. Automated Clinical Problem Detection from SOAP Notes using a Collaborative Multi-Agent LLM Architecture. 2025.

26. Nori H, Daswani M, Kelly C, et al. Sequential Diagnosis with Language Models. 2025.

27. Blondeel M, Codella N, Preston S, et al. Demo: Healthcare Agent Orchestrator (HAO) for Patient Summarization in Molecular Tumor Boards. 2025:arXiv:2509.06602. doi:10.48550/arXiv.2509.06602 Accessed September 01, 2025. https://ui.adsabs.harvard.edu/abs/2025arXiv250906602B

28. Wong C, Preston S, Liu Q, et al. Universal Abstraction: Harnessing Frontier Models to Structure Real-World Data at Scale. arXiv; 2025.

29. Benson R, Kenny C, Ashraf Ganjouei A, et al. Large Language Models in Population Oncology: A Contemporary Review on the Use of Large Language Models to Support Data Collection, Aggregation, and Analysis in Cancer Care and Research. JCO Clin Cancer Inform. Oct 2025;9:e2500112. doi:10.1200/CCI-25-00112

30. Hao Y, Qiu Z, Holmes J, et al. Large language model integrations in cancer decision-making: a systematic review and meta-analysis. NPJ Digit Med. Jul 17 2025;8(1):450. doi:10.1038/s41746-025-01824-7

31. Livingston L, Featherstone-Uwague A, Barry A, et al. Reproducible generative artificial intelligence evaluation for health care: a clinician-in-the-loop approach. JAMIA Open. Jun 2025;8(3):ooaf054. doi:10.1093/jamiaopen/ooaf054

32. Wells BJ, Nguyen HM, McWilliams A, et al. A practical framework for appropriate implementation and review of artificial intelligence (FAIR-AI) in healthcare. NPJ Digit Med. Aug 11 2025;8(1):514. doi:10.1038/s41746-025-01900-y

33. Harris PA, Taylor R, Thielke R, Payne J, Gonzalez N, Conde JG. Research electronic data capture (REDCap)--a metadata-driven methodology and workflow process for providing translational research informatics support. J Biomed Inform. Apr 2009;42(2):377–81. doi:10.1016/j.jbi.2008.08.010

34. Kaiser KN, Hughes AJ, Yang AD, et al. Accuracy and consistency of publicly available Large Language Models as clinical decision support tools for the management of colon cancer. J Surg Oncol. Oct 2024;130(5):1104–1110. doi:10.1002/jso.27821

35. Maynez J, Narayan S, Bohnet B, McDonald R. On Faithfulness and Factuality in Abstractive Summarization. arXiv; 2020.

36. Min S, Krishna K, Lyu X, et al. FActScore: Fine-grained Atomic Evaluation of Factual Precision in Long Form Text Generation. arXiv; 2023.

37. Ji Z, Lee N, Frieske R, et al. Survey of Hallucination in Natural Language Generation. ACM Comput Surv. 2023/12/31/ 2023;55(12):1–38. doi:10.1145/3571730

38. Tang L, Goyal T, Fabbri AR, et al. Understanding Factual Errors in Summarization: Errors, Summarizers, Datasets, Error Detectors. arXiv; 2023.

39. Gao Y, Xiong Y, Gao X, et al. Retrieval-Augmented Generation for Large Language Models: A Survey. arXiv; 2024.

40. Lewis P, Perez E, Piktus A, et al. Retrieval-Augmented Generation for Knowledge-Intensive NLP Tasks. arXiv; 2021.

41. Adler-Milstein J, Wang MD. The impact of transitioning from availability of outside records within electronic health records to integration of local and outside records within electronic health records. J Am Med Inform Assoc. Apr 1 2020;27(4):606–612. doi:10.1093/jamia/ocaa006

42. Rudin RS, Motala A, Goldzweig CL, Shekelle PG. Usage and effect of health information exchange: a systematic review. Ann Intern Med. Dec 2 2014;161(11):803–11. doi:10.7326/M14-0877

43. Li T, Zhang G, Do QD, Yue X, Chen W. Long-context LLMs Struggle with Long In-context Learning. arXivorg. 2024/04/02/ 2024;

44. Shi F, Chen X, Misra K, et al. Large Language Models Can Be Easily Distracted by Irrelevant Context. arXivorg. 2023/01/31/ 2023;

45. Liu NF, Lin K, Hewitt J, et al. Lost in the Middle: How Language Models Use Long Contexts. arXiv; 2023.

46. Adler-Milstein J, Holmgren AJ, Kralovec P, Worzala C, Searcy T, Patel V. Electronic health record adoption in US hospitals: the emergence of a digital “advanced use” divide. J Am Med Inform Assoc. Nov 1 2017;24(6):1142–1148. doi:10.1093/jamia/ocx080

47. Evans S, Seaman AT, Johnson EC, et al. Rural comprehensive cancer care: Qualitative analysis of current challenges and opportunities. J Rural Health. Sep 2024;40(4):634–644. doi:10.1111/jrh.12842

48. Poon EG, Lemak CH, Rojas JC, Guptill J, Classen D. Adoption of artificial intelligence in healthcare: survey of health system priorities, successes, and challenges. J Am Med Inform Assoc. Jul 1 2025;32(7):1093–1100. doi:10.1093/jamia/ocaf065

49. James JK, Maran T, Rice MP, et al. Experience With an Optical Character Recognition Search Application for Review of Outside Medical Records. Mayo Clin Proc Digit Health. Dec 2024;2(4):511–514. doi:10.1016/j.mcpdig.2024.08.001

50. Hsu E, Malagaris I, Kuo YF, Sultana R, Roberts K. Deep learning-based NLP data pipeline for EHR-scanned document information extraction. JAMIA Open. Jul 2022;5(2):ooac045. doi:10.1093/jamiaopen/ooac045

